# Twelve-Month Outcomes of Intrathecal Vesemnogene Lantuparvovec for Spinal Muscular Atrophy in Children Younger than 24 Months in Low- and Middle-Income Countries

**DOI:** 10.64898/2026.05.27.26354188

**Authors:** Lock Hock Ngu, Qingling Mo, Shiqi Li, Teck Hock Toh, Jia Ni Lee, Kok Chong Lim, Edi Setiawan Tehuteru, Rahmi Lestari, Chinnuwat Sanguansermsri, Hany Abueita, Samson Gwer, Li Li, Zhuowei Wang, Salman Kirmani, Julia X. Chen, Yilia Y. Cai, Nanette N. Zheng, Sherily Y. Yang, Paisley J. Liang, Yanqing Li, Mengdi Lu, Yanqiong Tang, Yeqing Li, Jason Z. Ye, Silk J. Shi, Jack F. Hong, Amy Y. Chen, Cole K. Zheng, Sanbin Wang, Teck-Onn Lim, Bruce T. Lahn, Austin T. Gao

## Abstract

**Introduction:** Spinal muscular atrophy (SMA) is a monogenic neuromuscular disease caused by mutations in the survival motor neuron 1 (*SMN1*) gene. Onasemnogene abeparvovec is a U.S. FDA-approved single-dose gene therapy for SMA. Both its intravenous formulation (Zolgensma, approximately USD 2.13 million per patient) and intrathecal formulation (Itvisma, around USD 2.59 million per patient) are prohibitively expensive, substantially limiting accessibility in low- and middle-income countries (LMICs). We conducted a clinical study of vesemnogene lantuparvovec, an alternative to onasemnogene abeparvovec developed for use in LMIC settings.

**Methods:** Sixteen patients with SMA, including 8 with type 1 SMA and 8 with type 2 SMA, received a single intrathecal administration of vesemnogene lantuparvovec. Eleven patients were treated with a low dose (1.5 × 10^14^ vg) and five with a high dose (3.0 × 10^14^ vg). The primary endpoints were safety and efficacy, assessed by changes from baseline in developmental gross motor milestones according to the World Health Organization criteria. Overall survival was primarily evaluated in type 1 SMA patients. This trial was registered with ClinicalTrials.gov NCT06288230.

**Results:** As of the March 2026 cutoff date, 15 of 16 treated patients had completed at least 12 months of follow-up after treatment, while the remaining one type 1 SMA patient died of disease progression at month 6 post-treatment. At 12 months post-treatment, among the surviving 7 patient with type 1 SMA, the median age was 21.6 months (range, 16.1 to 32.3 months). Among the 16 treated patients, the median age at diagnosis was 4.4 months (range, 0.0 to 18.0 months), and the median age at dosing was 10.7 months (range, 2.8 to 22.5 months). All patients experienced at least one AE. Thirty-one AESIs were reported in 13 patients, including hepatotoxicity, thrombocypenia-related events and cardiac events. No patient required prolonged prednisolone prophylaxis. SAEs, including pneumonia, lower respiratory tract infection, upper respiratory tract infection, and haemorrhagic diarrhoea, occurred in 5 of 8 (63%) patients with type 1 SMA and 2 of 8 (25%) patients with type 2 SMA. Two patients with type 1 SMA required invasive ventilation, and one of whom subsequently died.

At 12 months post-treatment, 11 of 16 treated patients (69%) gained at least one new WHO motor milestone versus baseline, including 3 type 1 and 8 type 2 SMA patients; one type 2 patient gained six WHO motor milestones and achieved independent walking.

**Conclusions:** In patients younger than 24 months of age with type 1 or type 2 SMA, a single intrathecal dose of vesemnogene lantuparvovec was safe and generally well tolerated and was associated with improvements in developmental gross motor milestones compared with outcomes observed among referred but untreated patients. Additional studies are required to further evaluate the long-term safety and efficacy of this gene therapy.

## Introduction

Spinal muscular atrophy (SMA) is a hereditary neuromuscular disease caused by mutations in the survival motor neuron 1 (*SMN1*) gene, leading to deficiency of SMN protein. With an estimated prevalence of approximately 1 in 14,694 live births,^[1]^ SMA represents one of the most common severe genetic disorders in infancy. Despite the availability of disease-modifying therapies such as nusinersen and risdiplam, SMA continues to contribute substantially to infant morbidity and mortality in low- and middle-income countries (LMICs), where the long-term costs and infrastructure requirements associated with lifelong treatment remain major barriers to care.

Gene replacement therapy with onasemnogene abeparvovec (Zolgensma, Novartis) offers a one-time treatment approach for SMA and may therefore be particularly advantageous in LMIC settings, where sustained access to chronic therapies can be difficult. Clinical studies have demonstrated pronounced efficacy in presymptomatic infants^[2,3]^ and in symptomatic children younger than six months of age.^[4–7]^ Therapeutic benefit has also been reported in patients up to 24 months old, although treatment responses appear to decline with increasing age at administration.^[8]^ Similar findings were reported in a recent study evaluating a single intrathecal administration of onasemnogene abeparvovec in older children.^[9,10]^

Zolgensma was approved by the U.S. FDA in 2019 for SMA patients younger than two years of age with the price of approximately USD 2.13 million per patient.^[11]^ In 2025, Itvisma also received the U.S. FDA approval for pediatric patients aged over 2 years and adults, at a reported cost of approximately USD 2.59 million per patient.^[12,13]^ The exceptionally high costs of these therapies greatly restrict their accessibility in LMICs. Development of lower-cost alternatives could therefore substantially reduce the disease burden associated with SMA in these regions. Vesemnogene lantuparvovec, an adeno-associated virus (AAV)-based gene therapy for SMA, was developed with this objective in mind. To maximize affordability, the therapy is intended to be offered at a price close to manufacturing costs. Its manufacturing technology may also be transferred to interested LMICs at minimal or no licensing fees to facilitate local gene therapy development and regulatory approval.

Vesemnogene lantuparvovec utilizes the vector construct rAAV9.OEE.h-SMN1. Similar to onasemnogene abeparvovec, it employs a nonreplicating recombinant adeno-associated virus serotype 9 (rAAV9) carrying a full-length, non-codon-optimized human *SMN1* transgene designed to restore SMN protein expression in motor neurons. The encoded SMN1 protein is structurally identical to endogenous wild-type human SMN1 protein. The expression cassette of vesemnogene lantuparvovec, flanked by inverted terminal repeat sequences, contains the human *SMN1* cDNA, an optimized promoter, an engineered post-transcriptional regulatory element, and a bovine growth hormone (BGH) polyadenylation sequence. By comparison, the expression cassette of onasemnogene abeparvovec-xioi includes the chicken β-actin promoter, simian virus 40 intron, and BGH polyadenylation sequence.

Vesemnogene lantuparvovec has been evaluated in a clinical study conducted in China (ClinicalTrials.gov identifier: NCT06288230). Here, we present preliminary safety and efficacy findings from all treated children as of March 2026.

## Methods

### Study design and oversight

This prospective, single-center, dose-escalation study was carried out in Kunming, China. The trial was performed in compliance with the principles outlined in the Declaration of Helsinki. Approval of the study protocol was obtained from the institutional review board of the participating center, and written informed consent was provided by all parents or legal guardians prior to enrollment. The study was registered at ClinicalTrials.gov (NCT06288230).

### Study patients

Eligible participants were children aged 24 months or younger with genetically confirmed SMA caused by biallelic *SMN1* deletion and lacking the genetic modifier variant c.859G>C.^[14]^ Both symptomatic and presymptomatic patients were eligible for enrollment. Key exclusion criteria included contraindications to lumbar puncture or intrathecal administration, anti-AAV9 antibody titers greater than 1:50 measured by ELISA, requirement for invasive ventilatory support (tracheostomy with positive-pressure ventilation) or noninvasive ventilatory support for at least 12 hours per day, severe scoliosis or joint contractures, active respiratory or systemic infection, abnormal hepatic function defined as alanine aminotransferase (ALT) or aspartate aminotransferase (AST) levels exceeding twice the upper limit of normal, and any additional coexisting medical condition considered by the investigators to increase the potential risk associated with gene replacement therapy. Participants meeting eligibility criteria received a single treatment at either a low dose (1.5 × 10^14^ vg) or a high dose (3.0 × 10^14^ vg).

### Study treatment

All participants received study treatment according to a standardized protocol. Vesemnogene lantuparvovec was administered as a single intrathecal dose under sedation or general anesthesia. After injection, patients were maintained in the Trendelenburg position for 15 minutes to facilitate vector distribution toward the cervical spinal cord and brain regions. All patients remained hospitalized for observation for a minimum of 48 hours after treatment before discharge. Prophylactic oral prednisolone at a dose of 1 mg per kilogram per day was started one day before administration of intrathecal vesemnogene lantuparvovec and maintained for four weeks. If liver function tests remained within the normal range, prednisolone was subsequently tapered over a period of two to four weeks.

### Study assessments

Safety evaluations included monitoring for adverse events (AEs), serious adverse events (SAEs), and adverse events of special interest (AESIs), together with assessments of hematologic parameters, blood chemistry profiles, and findings from physical examinations. AEs were coded according to the preferred terminology of the Medical Dictionary for Regulatory Activities (MedDRA).

The primary efficacy endpoint was the change from baseline in developmental gross motor milestones as defined by the World Health Organization (WHO), including sitting without support, standing with assistance, crawling on hands and knees, walking with assistance, standing independently, and independent walking.^[15]^ Motor milestone assessments were recorded on video and underwent centralized review. Overall survival was primarily evaluated in type 1 SMA patients. Secondary efficacy endpoints included the need for noninvasive ventilatory support and nutritional support via nasogastric or gastrostomy tube feeding.

### Statistical analysis

Safety and efficacy were evaluated in the intention-to-treat population, which included all patients who received a single dose of vesemnogene lantuparvovec. Safety analysis was evaluated based on the incidence of AEs, SAEs, and AESIs. Efficacy analyses assessed changes in the distribution of WHO motor milestones from baseline to post-treatment, individual trajectories of motor milestone development following treatment, and the proportion of patients achieving at least one WHO motor milestone.

## Results

The first patient underwent treatment in May 2024. Following the absence of significant safety concerns in the initial three treated patients, enrollment was subsequently expanded to include referrals from LMICs worldwide. Since October 2024, a substantial number of patients were referred for screening and eligibility assessment. Among 128 screened patients, 102 were excluded because they had previously been referred for palliative care (n = 16), or were positive for anti-AAV antibodies (n = 4), or were older than 24 months of age (n = 71), or failed to meet the eligibility criteria (n = 11). A total of 26 patients met the eligibility criteria. Treatment capacity was limited by the availability of doses from the clinical trial manufacturing batch. By March 2026, 16 eligible patients had received treatment, and 5 patients died while awaiting therapy and another 5 remained on the waiting list. All treated patients completed 12 months of follow-up after treatment, with no study discontinuations other than one patient who died during the follow-up period.

**Figure 1.**
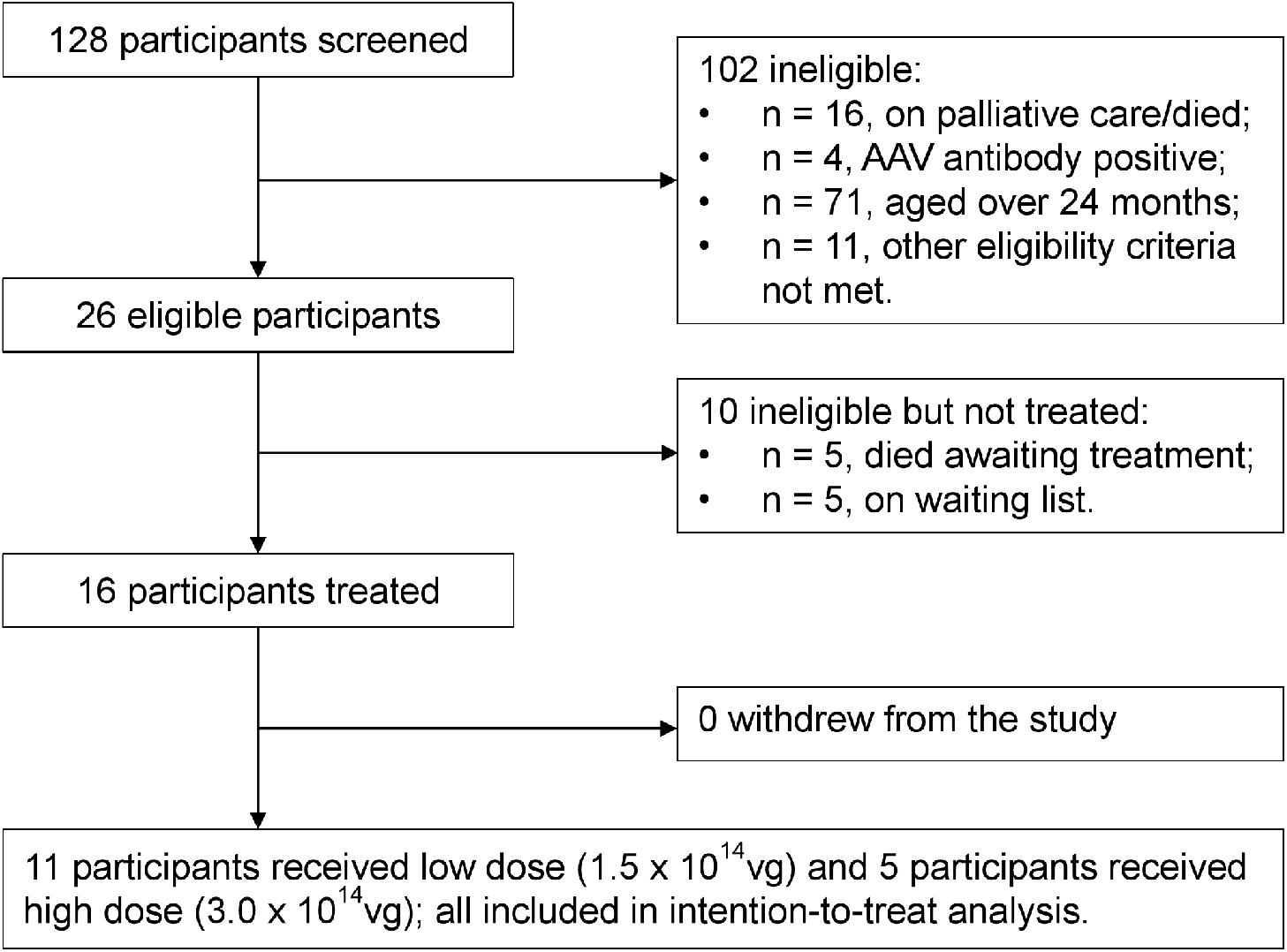
Trial Profile.

Among the 16 treated patients, five were from Vietnam, four from Pakistan, two from Palestine, and one each from Malaysia, Thailand, Indonesia, India, and Kenya. Baseline patient characteristics are summarized in **Table 1**. The median age at treatment administration was 10.7 months, with a range of 2.8 to 22.5 months. Eight patients carried two copies of *SMN2*, and the remaining eight carried three copies. Eight patients were classified as type 1 SMA, with symptom onset before 6 months of age, diagnosis before 12 months of age, and two *SMN2* copies. The other eight patients had type 2 SMA. Before enrollment, 11 patients had been treated with risdiplam, while none had received nusinersen. At baseline, most patients were unable to sit independently; three could sit without support and two could stand with assistance. Among patients with type 1 SMA, only one was able to sit unsupported at baseline. Of the eight patients with type 1 SMA, one required nasogastric tube feeding and another required gastrostomy tube feeding. One patient required invasive ventilatory support, and three patients had previously required intermittent bilevel positive airway pressure (BiPAP) support during respiratory infections. None of the patients with type 2 SMA required tube feeding or BiPAP support at baseline.

**Table 1.**
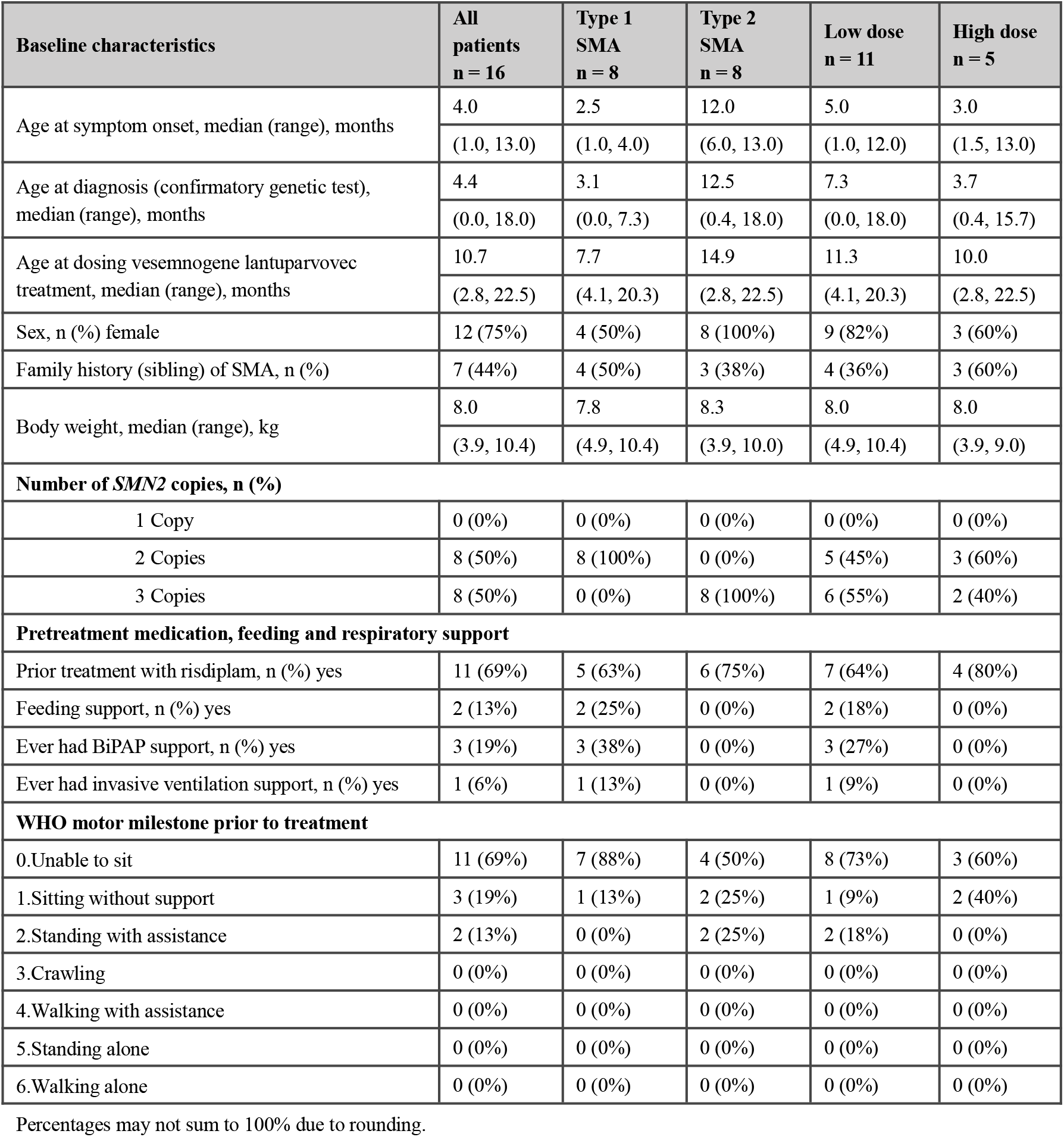
Baseline characteristics of patients.

### Safety results

As of March 2026, AEs had been reported in all treated patients (**Table 2**). SAEs occurred in 7 patients, including 6 in the low-dose cohort and one in the high-dose cohort. Pneumonia was the most commonly reported SAE, occurring in 4 patients (25%). And lower respiratory tract infection was reported in 2 patients (13%). Other SAEs included haemorrhagic diarrhoea and upper respiratory tract infection, each reported in one patient (6%). Four respiratory tract infections SAEs were associated with hypoxemia requiring ventilatory support, including two patients who subsequently required invasive ventilation. One patient died 6 months after treatment. Investigators considered these complications to be consistent with the natural progression of type 1 SMA. No patients with type 2 SMA required ventilatory support during follow-up.

**Table 2.** Overview of AEs.

| **Patients with AEs, n (%)** | **Total (n = 16)** |
| --- | --- |
| Any AE* | 16 (100%) |
| **SAEs** | **7 (44%)** |
| Pneumonia | 4 (25%) |
| Other lower respiratory tract infection | 2 (13%) |
| Upper respiratory tract infection | 1 (6%) |
| Haemorrhagic diarrhoea | 1 (6%) |
| Any AE leading to death | 1 (6%) |
| Any AE leading to study discontinuation | 0 (0%) |
| **AESIs** | **13 (81%)** |
| Hepatotoxicity | 12 (75%) |
| AST increased | 11 (69%) |
| ALT increased | 3 (19%) |
| ALP increased | 2 (13%) |
| γ-glutamyltransferase increased | 1 (6%) |
| Thrombocytopenia and related AEs | 6 (38%) |
| Blood fibrinogen decreased | 4 (25%) |
| Anaemia | 4 (25%) |
| Diarrhoea haemorrhagic | 1 (6%) |
| Thrombocytopenia | 0 (0%) |
| Cardiac AEs | 2 (13%) |
| Troponin I increased | 1 (6%) |
| Troponin T increased | 1 (6%) |
| TMA and related AEs | 0 (0%) |
| Sensory abnormalities | 0 (0%) |
| **Most frequent AEs (≥25%)** | **15 (94%)** |
| AST increased | 11 (69%) |
| Pyrexia | 6 (38%) |
| Thrombocytosis | 6 (38%) |
| Pneumonia | 6 (38%) |
| Low density lipoprotein increased | 5 (31%) |
| Lymphocyte count increased | 4 (25%) |
| Anaemia | 4 (25%) |
| Blood lactate dehydrogenase increased | 4 (25%) |
| Blood fibrinogen decreased | 4 (25%) |

Note: Percentages may not sum to 100% due to rounding. A participant with multiple occurrences of an AE for a preferred term is counted only once in each specific category (MedDRA version 28.1).
\* One hundred and seventeen AEs occurred among 16 patients; all patients have at least one event.

AESIs were assessed, including hepatotoxicity, thrombocytopenia-related events, cardiac events, thrombotic microangiopathy (TMA), and sensory abnormalities (**Table 2**). A total of 19 hepatotoxicity-related events occurred in 12 of the 16 patients (75%). Liver enzyme elevations included increased AST in 11 patients (69%), ALT in 3 patients (19%), alkaline phosphatase (ALP) in 2 patients (13%), and γ-glutamyltransferase in one patient (6%). Ten thrombocytopenia-related events occurred in 6 of the 16 patients (38%). Abnormal haematological findings included decreased fibrinogen in 4 patients (25%), anaemia in 4 patients (25%), and haemorrhagic diarrhoea in one patient (6%). Cardiac AESIs were reported in 2 patients (12%), including elevated troponin I in one patient and elevated troponin T in the other. No clinical evidence of TMA or sensory abnormalities was identified; however, routine electrophysiological testing was not performed in these young children.

Elevations in AST, ALT, and troponin I levels and anaemia were considered related to study treatment. All aminotransferase elevations resolved spontaneously. All patients received prophylactic prednisolone, and none required corticosteroid treatment beyond the planned 4-week course.

The most frequently observed AEs are summarized in **Table 2**. Increased AST was the most common event, occurring in 11 patients (69%). This was followed by pyrexia, thrombocytosis, and pneumonia, each reported in 6 patients (38%).

Additional events included endoscopic placement of nasogastric or gastrostomy feeding tubes to reduce aspiration risk and provide nutritional support (6 events, 38%). Of these 6 patients, two were already receiving feeding support at baseline. All such interventions occurred in patients with type 1 SMA, including five patients in the low-dose cohort and one patient in the high-dose cohort.

### Efficacy results

All parents reported noticeable improvements in their children’s motor function within days to weeks following treatment with vesemnogene lantuparvovec. Newly observed abilities reported by caregivers included visually tracking moving objects, grasping objects with the hands, unsupported head control, and rolling from the back to the side.

Changes in the distribution of WHO motor milestones from baseline to post-treatment follow-up are summarized in **Figure 2** and **Table 3**. By 12 months after treatment, 11 of 16 patients (69%) gained at least one new WHO motor milestone. Five patients gained one milestone, five gained two milestones, and one gained six milestones.

**Table 3.** WHO motor milestones at baseline and achieved post-treatment.

| WHO motor milestone | BL | M1 | M3 | M6 | M9 | M12 |
| --- | --- | --- | --- | --- | --- | --- |
|  | n = 16 | n = 16 | n = 16 | n = 16* | n = 16* | n = 16* |
| Unable to sit | 11 (69%) | 8 (50%) | 9 (56%) | 7 (44%) | 6 (38%) | 3 (19%) |
| Sitting without support | 3 (19%) | 6 (38%) | 4 (25%) | 4 (25%) | 5 (31%) | 6 (38%) |
| Standing with assistance | 2 (13%) | 2 (13%) | 0 (0%) | 1 (6%) | 1 (6%) | 1 (6%) |
| Crawling | 0 (0%) | 0 (0%) | 3 (19%) | 1 (6%) | 0 (0%) | 2 (13%) |
| Walking with assistance | 0 (0%) | 0 (0%) | 0 (0%) | 2 (13%) | 2 (13%) | 2 (13%) |
| Standing alone | 0 (0%) | 0 (0%) | 0 (0%) | 0 (0%) | 0 (0%) | 0 (0%) |
| Walking alone | 0 (0%) | 0 (0%) | 0 (0%) | 0 (0%) | 1 (6%) | 1 (6%) |

BL: Baseline, M1: 1 months post-treatment, M3: 3 months post-treatment, M6: 6 months post-treatment, M9: 9 months post-treatment, M12: 12 months post-treatment.
\*: One patient died at 6-month post-treatment.

**Figure 2.**
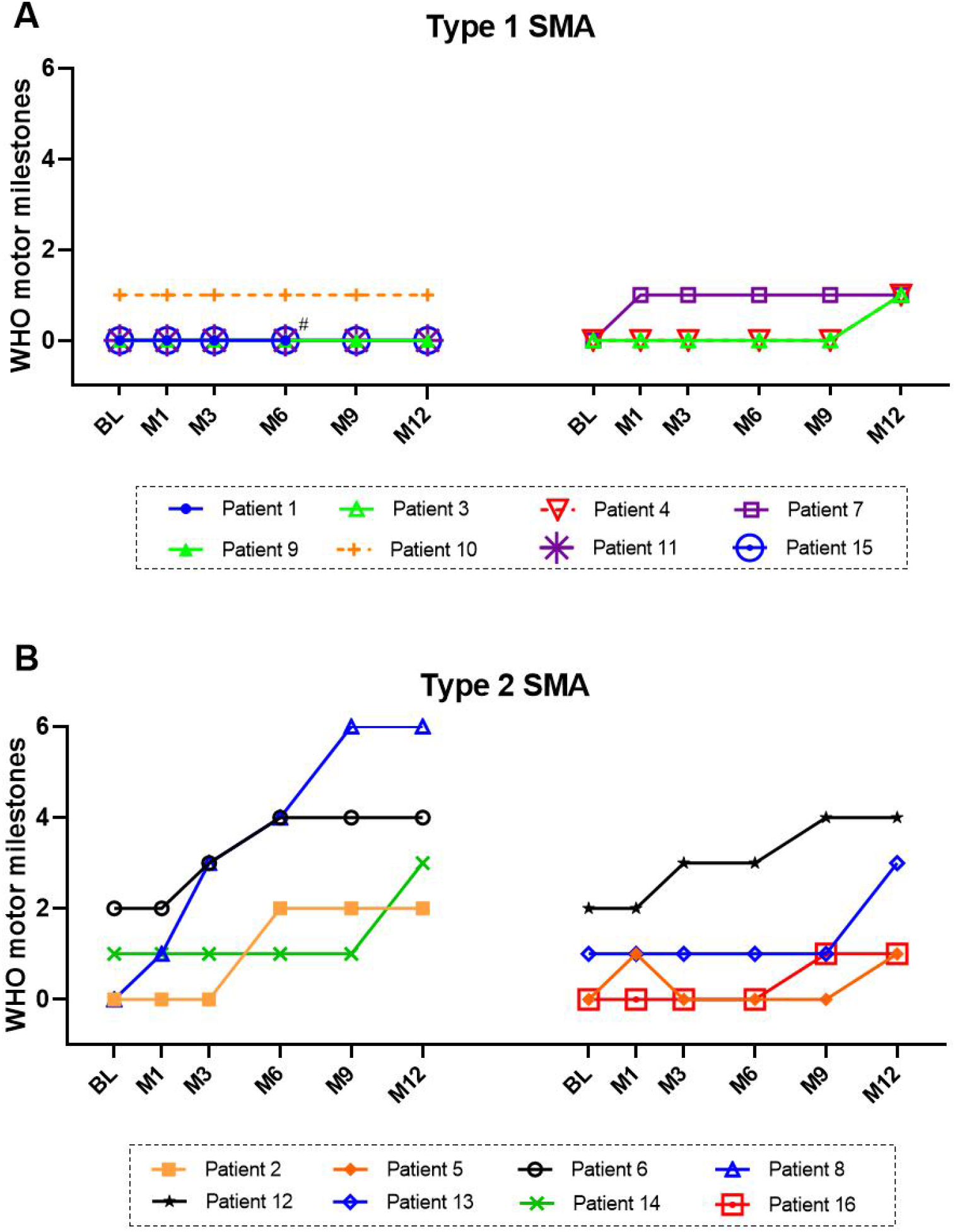
Trajectory of WHO motor milestones achieved post-treatment among type 1 patients (A) and type 2 patients (B). Abbreviations: BL: Baseline, M1: 1 months post-treatment, M3: 3 months post-treatment, M6: 6 months post-treatment, M9: 9 months post-treatment, M12: 12 months post-treatment. #, patient 1 died in M6.

By the data cutoff, 7 of 8 type 1 SMA patients were alive without permanent ventilation continuing in the trail ranged in age from 16.1 to 32.3 months (median age, 21.6 months). During follow-up, 6 of 8 type 1 SMA patients (75%) required feeding support and 4 (50%) required non-invasive ventilation support during respiratory infections. Among the four patients, one died at 6 months after treatment following progression to invasive ventilation; one needed invasive ventilation at 3 months post-treatment, with subsequent recovery of spontaneous unassisted ventilation. In addition to survival, 3 of 8 type 1 SMA patients gained a new WHO motor milestone at Month 12 versus baseline; one patient achieved this milestone as early as month 1 and sustained the improvement through Month 12 (**Table 4**).

**Table 4.** WHO motor milestones in type 1 SMA patients.

| WHO motor milestone | BL | M1 | M3 | M6 | M9 | M12 |
| --- | --- | --- | --- | --- | --- | --- |
|  | n = 8 | n = 8 | n = 8 | n = 8* | n = 8* | n = 8* |
| Unable to sit | 7 (88%) | 6 (75%) | 6 (75%) | 5 (63%) | 5 (63%) | 3 (38%) |
| Sitting without support | 1 (13%) | 2 (25%) | 2 (25%) | 2 (25%) | 2 (25%) | 4 (50%) |
| Standing with assistance | 0 (0%) | 0 (0%) | 0 (0%) | 0 (0%) | 0 (0%) | 0 (0%) |
| Crawling | 0 (0%) | 0 (0%) | 0 (0%) | 0 (0%) | 0 (0%) | 0 (0%) |
| Walking with assistance | 0 (0%) | 0 (0%) | 0 (0%) | 0 (0%) | 0 (0%) | 0 (0%) |
| Standing alone | 0 (0%) | 0 (0%) | 0 (0%) | 0 (0%) | 0 (0%) | 0 (0%) |
| Walking alone | 0 (0%) | 0 (0%) | 0 (0%) | 0 (0%) | 0 (0%) | 0 (0%) |

All eight patients with type 2 SMA gained at least one new WHO motor milestone at 12 months after treatment. Two patients gained one milestone, advancing from inability to sit to sitting without support. Five patients achieved gains of two milestones: one advanced from inability to sit to standing with assistance, two advanced from sitting independently to crawling, and two advanced from standing with assistance to walking with assistance. One patient gained six milestones and became able to walk independently (**Table 5**). No patients with type 2 SMA required ventilatory or feeding support during follow-up.

**Table 5.**
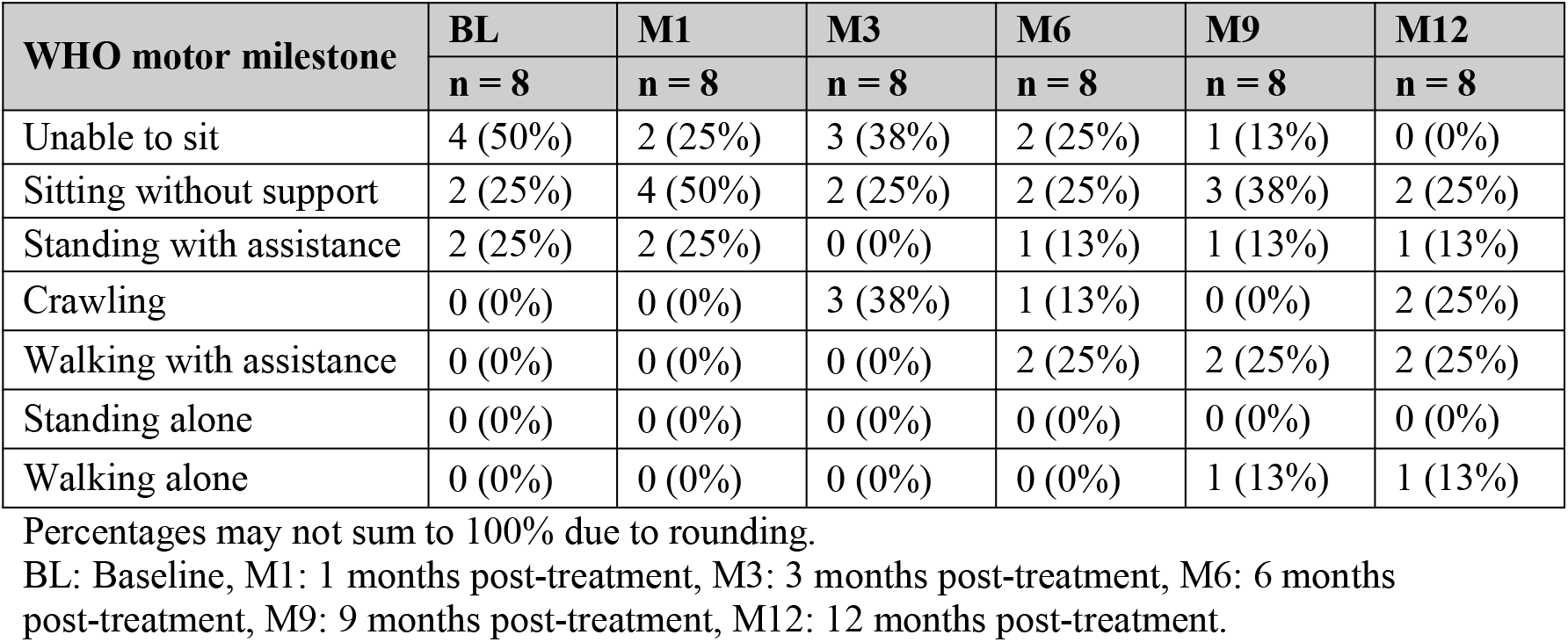
WHO motor milestones in type 2 SMA patients.

The distribution of WHO motor milestone outcomes in the low-dose (1.5 × 10^14^ vg) and high-dose (3.0 × 10^14^ vg) groups is presented in **Table6** and **Table 7**. At 12 months post-treatment, no significant advantage in WHO motor milestone outcomes was observed in the high-dose cohort compared with the low-dose cohort.

**Table 6.** WHO motor milestones in low dose (1.5 × 10^14^ vg)

| WHO motor milestone | BL | M1 | M3 | M6 | M9 | M12 |
| --- | --- | --- | --- | --- | --- | --- |
|  | n = 11 | n = 11 | n = 11 | n = 11* | n = 11* | n = 11* |
| Unable to sit | 8 (73%) | 5 (45%) | 6 (55%) | 4 (36%) | 4 (36%) | 1 (9%) |
| Sitting without support | 1 (9%) | 4 (36%) | 2 (18%) | 2 (18%) | 2 (18%) | 4 (36%) |
| Standing with assistance | 2 (18%) | 2 (18%) | 0 (0%) | 1 (9%) | 1 (9%) | 1 (9%) |
| Crawling | 0 (0%) | 0 (0%) | 3 (27%) | 1 (9%) | 0 (0%) | 1 (9%) |
| Walking with assistance | 0 (0%) | 0 (0%) | 0 (0%) | 2 (18%) | 2 (18%) | 2 (18%) |
| Standing alone | 0 (0%) | 0 (0%) | 0 (0%) | 0 (0%) | 0 (0%) | 0 (0%) |
| Walking alone | 0 (0%) | 0 (0%) | 0 (0%) | 0 (0%) | 1 (9%) | 1 (9%) |

BL: Baseline, M1: 1 months post-treatment, M3: 3 months post-treatment, M6: 6 months post-treatment, M9: 9 months post-treatment, M12: 12 months post-treatment.
\*: one patient died at 6-month post-treatment.

**Table 7.**
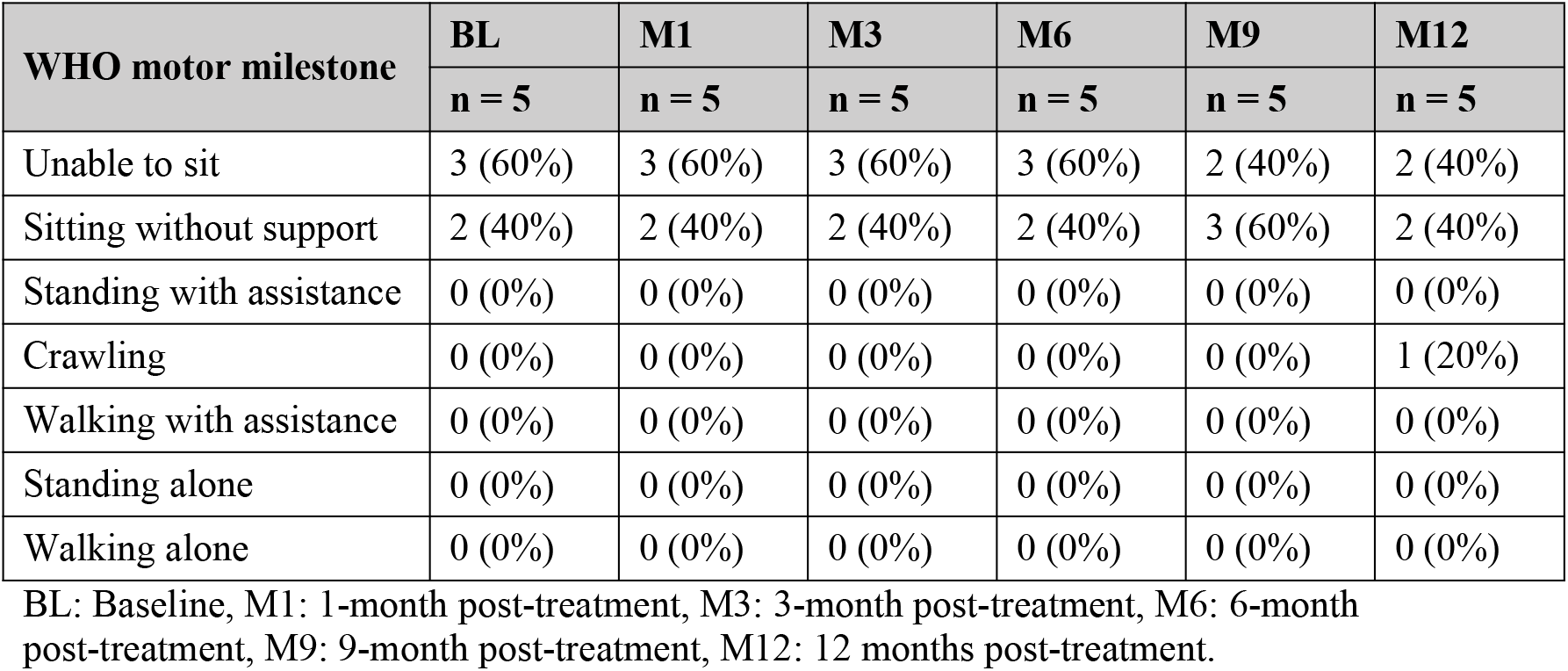
WHO motor milestones at baseline and achieved post-treatment of high dose (3.0 × 10^14^ vg)

## Discussion

We conducted a clinical evaluation of vesemnogene lantuparvovec in children with SMA enrolled from multiple LMICs across diverse geographic regions and patient populations.

Based on prior experience with AAV-mediated gene therapy, acute AEs are anticipated to occur predominantly during the early post-treatment period, with immune-mediated hepatic and cardiac toxicities particular concern. Consistent with previous reports of AAV-based SMA gene therapies, transient elevations of transaminases were observed in most patients within the first month after treatment with vesemnogene lantuparvovec. However, unlike the hepatic events described following intravenous administration of onasemnogene abeparvovec, the transaminase elevations observed in this study occurred as isolated laboratory abnormalities without accompanying clinical signs or symptoms and resolved spontaneously in all affected patients. No patient required extension of prednisolone prophylaxis beyond the planned treatment period. Elevated troponin I was observed in only one patient with no abnormal clinical manifestations observed. Overall, single-dose intrathecal administration of vesemnogene lantuparvovec demonstrated a favorable safety and tolerability profile, consistent with findings from the previous study of intrathecal onasemnogene abeparvovec administration.^[9]^

With respect to efficacy, the current analysis remains limited by the small number of patients and duration of follow-up, with data available for only 15 patients through 12 months after treatment. Nevertheless, by 12 months post-treatment, 7 of 8 (88%) treated type 1 SMA patients survived and 11 of 16 treated patients (69%) achieved gains of at least one WHO motor milestone. Notably, one patient improved by six WHO motor milestones by 9 months after treatment and maintained the ability to walk independently through the 12-month follow-up period.

However, interpretation of these findings should be cautious given the single-arm design of the study. In the absence of a comparable control group, potential confounding factors cannot be excluded, making it difficult to determine to what extent the observed motor improvements were attributable to normal developmental maturation versus the therapeutic effect of vesemnogene lantuparvovec.

A randomized placebo-controlled design would have been methodologically preferable but was considered ethically difficult, even in LMIC settings where palliative care remains the standard approach for most infants and young children with type 1 SMA.

Establishing a suitable natural-history comparator cohort in this context was also not feasible. Patients enrolled in this study were referred from multiple LMICs with considerable heterogeneity in local standards of care. In many of these settings, access to genetic testing for SMA remains limited, and infants presenting with hypotonia, progressive weakness, and respiratory failure are often referred directly to palliative care without molecular confirmation of the diagnosis. In addition, access to charity-supported risdiplam treatment varies substantially across countries. Consequently, patients enrolled in this trial represented a highly selected subgroup of the broader SMA population by virtue of having received a confirmed diagnosis and access to referral pathways; notably, 69% had previously received risdiplam.

The most clinically comparable untreated patients were those referred for the trial but ultimately not treated. Two such groups were identified. The first consisted of patients with type 1 SMA who were excluded because of positive anti-AAV antibody testing. Among three such patients, two progressed to respiratory failure requiring invasive ventilation within 3 months after referral. The second group comprised patients with type 1 SMA who remained on the treatment waiting list. Treatment delays were unavoidable and included the time required for families and physicians to consider participation, delays associated with genetic and anti-AAV antibody testing requiring shipment of samples to a central laboratory, and delays related to obtaining passports and visas for travel to China. Among these seven patients, five died within 6 months after referral.

The rapidly progressive clinical deterioration observed among referred but untreated patients contrasted with outcomes in treated patients. At 12 months post-treatment, all gained at least one WHO motor milestone, including one patient who gained six milestones and achieved independent walking. These observations are consistent with previous studies of AAV-based gene therapy demonstrating rapid onset of clinical benefit,^[9]^ including in patients previously treated with disease-modifying therapies.^[10]^ Comparable rapid and marked motor improvements have not been observed among patients on the waiting list, including those receiving risdiplam.

This study has several important limitations. First, as discussed above, the lack of a randomized control group and reliance on external comparators introduce the potential for confounding bias. This limitation is common in interventional studies involving rare diseases, particularly when randomized controlled designs are not practically achievable or are considered ethically challenging.^[16]^ Second, the sample size remains relatively small for a clinical study. Additional later-phase studies are planned in LMICs to support local regulatory approval, and technology transfer of vesemnogene lantuparvovec to participating LMICs is currently in progress. Third, follow-up duration is presently limited to 12 months after treatment. Further evaluation of long-term safety and efficacy will require observation of larger patient cohorts over extended follow-up periods.

In conclusion, this study provides preliminary evidence that intrathecal administration of vesemnogene lantuparvovec, a novel AAV9-based gene therapy, is safe and generally well tolerated in young children with type 1 and type 2 SMA from LMIC settings. Although the cohort size was limited and follow-up duration remains relatively short, early improvements in WHO motor milestones, particularly among patients with type 2 SMA, are encouraging. Importantly, these clinical benefits were observed in a population with limited access to standard disease-modifying therapies and in healthcare settings where gene therapy has historically remained inaccessible because of cost constraints.

Interpretation of efficacy outcomes is limited by the absence of a randomized controlled group and the heterogeneity of the study population. Nevertheless, the marked contrast between treated patients and referred but untreated patients, many of whom experienced rapid disease progression and high mortality, highlights the potential therapeutic value of vesemnogene lantuparvovec.

These findings emphasize the urgent need for affordable and scalable gene therapy strategies tailored to LMIC settings. Larger studies with longer follow-up durations and locally relevant patient registries will be necessary to confirm long-term safety and efficacy, support regulatory approval, and facilitate broader clinical implementation. Ongoing efforts involving technology transfer and local manufacturing may represent important steps toward sustainable SMA treatment programs and more equitable global access to gene therapy.

## Data Availability

All data produced in the present study are available upon reasonable request to the authors

